# Association between maternal history of psychiatric disorders and the risk of cardiovascular disease in offspring up to early middle-age

**DOI:** 10.64898/2026.06.01.26354560

**Authors:** Fen Yang, Tai Ren, Imre Janszky, Hui Wang, Fei Li, Jiong Li, Krisztina D. László

## Abstract

**Objective:** To evaluate the associations between maternal history of psychiatric disorders and the risk cardiovascular disease (CVD) in offspring.

**Design:** Population based cohort study.

**Setting:** Nationwide health registers in Sweden.

**Participants:** All 4 171 005 liveborn singletons in Sweden from 1973 to 2014. Follow-up started at birth and ended until the first diagnosis of CVD, death, emigration, or December 31^st^, 2023, whichever occurred first.

**Exposures for observational studies:** Maternal psychiatric disorders diagnosed before delivery (n=208,680, 5.0%).

**Main outcome measures:** The primary outcome was the first diagnosis of CVD in offspring, identified through hospital registers. Additional outcomes included specific CVD subtypes. To address potential familial confounding, a cousin comparison was performed, comparing the risk of CVD in offspring born to mothers who were biological sisters. Mediation analyses examined the roles of congenital heart disease, small for gestational age, and preterm birth.

**Results:** During up to 51 years of follow-up, 307 596 (7.4%) offspring had a diagnosis of CVD. Maternal history of psychiatric disorders was associated with a higher risk of overall CVD both in the full cohort (hazard ratio 1.19, 95% confidence interval 1.17 to 1.21) and the cousin-comparison cohort (n=1 577 113; 1.08, 1.03 to 1.13). In disease-specific analyses, a prominent association with heart failure was robustly observed in both the full cohort (1.59, 1.37 to 1.85) and the cousin comparison cohort (1.51, 1.06 to 2.17). Mediation analyses indicated that congenital heart disease mediated 9.5% of the association between maternal psychiatric disorders and offspring CVD risk. Preterm birth and small for gestational age contributed minimally (<3%) to the observed associations.

**Conclusions:** Maternal history of psychiatric disorders was associated with an increased risk of CVD up to early middle-age in offspring. Congenital heart disease partly mediated this association.

## Introduction

Cardiovascular diseases (CVD) is the leading cause of morbidity and mortality worldwide.^1^ Though traditionally recognized as a morbidity of later adulthood, CVD incidence and prevalence have been increasing among adolescents and young adults.^2^ The etiology of CVD in these younger age group is likely to differ from that in older adults, with mounting evidence suggesting a role of fetal developmental programming and *in utero* exposures in their development.^3,4^ Of particular concern is the high prevalence of psychiatric disorders among women of childbearing age, with more than half of the psychiatric disorders globally presenting before the age of 25 years.^5^ However, empirical evidence investigating the association between maternal psychiatric disorders and offspring CVD risk remains notably scarce.

Psychiatric disorders may alter the intrauterine environment through the dysregulation of the hypothalamic-pituitary-adrenal axis,^6,7^ which may impact fetal cardiovascular development and cardiometabolic programming, potentially predisposing the offspring to increased CVD risk.^8,9^ Furthermore, a maternal history of psychiatric illness is frequently accompanied by adverse health behaviors during pregnancy.^10^ These behavioral factors, acting in concert with underlying biological mechanisms, might contribute to a higher incidence of congenital heart disease and adverse birth outcomes, which may further influence the risk of CVD in offspring. ^11–16^ Increasing evidence suggests that maternal depression and stress during pregnancy are associated with adverse cardiometabolic profiles in offspring.^17,18^ However, to our knowledge, no previous study has examined comprehensively the full spectrum of maternal psychiatric disorders in relation to CVD, nor the mechanisms underlying such a potential association.

In this population-based study, we leveraged Swedish nationwide data to analyze the association between maternal history of psychiatric disorders and offspring CVD risk from childhood to early adulthood. Furthermore, we used mediation analysis to test the hypothesis that this association can partially be explained by an increased risk of adverse birth outcomes and offspring congenital heart disease associated with maternal psychiatric disorders.

## Methods

### Study design and population

Each Swedish resident is assigned a unique personal identification number at birth, which enables individual-level linkage across multiple nationwide registers. The registers used in the present study are described in eTable 1.^19^ The study population included all liveborn singletons recorded in the Swedish Medical Birth Register (MBR) between 1973 and 2014.^20^ After excluding births with missing maternal personal identification number, the final cohort comprised 4,171,005 mother-child pairs. We followed each offspring from birth until the first diagnosis of CVD, death, emigration (available until December 31, 2014), or December 31^st^, 2023, whichever occurred first.

### Measures

#### Exposure

Offspring were classified as exposed if their mothers had a diagnosis of any psychiatric disorder prior to the index delivery; otherwise, they were considered unexposed. Maternal diagnoses of psychiatric disorders were obtained from the MBR and from the Patient Register^21^, using the International Statistical Classification of Diseases and Related Health Problems (ICD) codes shown in in eTable 2. The Patient Register was established in 1964. Its coverage with respect to psychiatric inpatient care was complete in all but five of the 25 regions during 1973-1987, when coverage became nationwide. Coverage of data from specialised non-private outpatient care became nationwide in 2001^22^. Psychiatric disorders were further categorized into the following 11 subtypes using a previously established algorithm^23^: 1) Organic psychoses, 2) Schizophrenia and other non-affective psychoses, 3) Affective disorders, 4) Neurotic disorders, 5) Stress-related disorders, 6) Personality disorder, 7) Attention deficit hyperactivity disorders, 8) Autism spectrum disorders, 9) Disorders related to psychoactive substance use, 10) Intellectual disability, 11) Somatoform disorders. The diagnostic codes used to identify these conditions are provided in eTable 2.

#### Outcome

Information on the first diagnosis of overall CVD was extracted from the Patient Register and from the Cause of Death Register^24^ using the ICD codes in eTable 2. In addition to overall CVD, we examined several common and severe CVD subtypes, including ischemic heart disease (with a separate analysis for acute myocardial infarction), stroke (further classified into hemorrhagic and ischemic strokes), heart failure, atrial fibrillation (including atrial flutter), hypertensive disease, and peripheral artery disease. Overall CVD, ischemic heart disease, stroke, and heart failure were identified using the primary diagnosis from the Patient Register and underlying cause of death using the Cause of Death Register. The remaining CVD types were identified based on either primary or secondary diagnoses in the Patient Register.

#### Covariates

We retrieved information on maternal (i.e., country of origin, marital status, highest level of education, age at delivery, parity, body-mass index (BMI) and smoking in early pregnancy, hypertensive disease and diabetes before or during the index pregnancy, and family history of CVD), paternal (any psychiatric disorder diagnosed before the index birth), and offspring characteristics (i.e., birth year, sex, gestational age, birth weight, and the diagnosis of congenital heart disease during the follow-up period) as described in eAppendix 1 in Supplement 1.

### Statistical analyses

We visualised the cumulative incidence of CVDs in offspring exposed and unexposed to maternal psychiatric disorders using Kaplan-Meier curves. We calculated crude incidence rates of overall CVD and its major subtypes by exposure status, defined as the number of incident cases per 10,000 person-years of follow-up.

In the full cohort, we estimated hazard ratios (HRs) and 95% confidence intervals (CIs) for overall CVD and its major subtypes according to maternal psychiatric disorder using Cox proportional hazards models, with attained age as the underlying time scale. We assessed the proportional hazards assumption using Schoenfeld residuals; no strong violations of the assumption were detected. The model was adjusted for offspring’s sex and birth year, as well as for maternal country of origin, parity, age, education, marital status at the time of birth, maternal family history of CVD prior to the index birth, maternal hypertensive disorders and diabetes before delivery.^3,25–27^ Analyses were further conducted for each of the 11 specific types of maternal psychiatric disorders, as well as each of the 9 types of CVD outcomes.

To further account for unmeasured confounding due to shared familial genetic and early-life environmental factors, we additionally utilized a cousin-comparison design: we identified offspring of women and their biological sisters from the full cohort and constructed a sub-cohort consisting of these cousin groups (N=1,577,113). In the cousin comparison cohort, we fitted conditional Cox proportional hazards models, assigning a separate baseline hazard function to each stratum of cousin groups, thereby enabling comparisons within extended families. The models were adjusted for the same set of covariates as in the main analysis (Model 3), with the exception of family history of CVD, which is inherently controlled for by the design.

### Mediation analysis

Maternal psychiatric disorders have been associated with preterm birth, fetal growth restriction, and congenital heart disease,^14–16^ which, in turn, have been linked to an increased risk of CVD later in life.^11–13^ Therefore, we hypothesized that these adverse birth outcomes and congenital heart disease may lie on the causal pathway between maternal psychiatric disorders and the risk of CVD in offspring. Counterfactual mediation analysis was conducted to assess the mediating roles of preterm birth (birth before 37 completed gestational weeks), small for gestational age (a proxy for fetal growth restriction, defined as birth weight below the 10th percentile, with reference to the Swedish sex-specific curve for normal fetal growth^28^), and congenital heart disease.^29^ In brief, the total effect of maternal psychiatric disorders on offspring CVD, measured by HRs, was decomposed into a direct effect (DE) and a mediated (indirect) effect (ME). Within this framework, exposure-mediator interactions were explicitly modelled and incorporated into the estimation of the indirect effect. The proportion mediated by each factor was calculated using the formula:

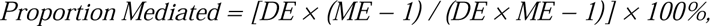

where 0% indicates no mediation and 100% indicates complete mediation (i.e., no direct effect). Proportion mediated was reported if DE and ME were in the same direction.

### Sensitivity analyses

To account for paternal factors, we additionally adjusted for paternal psychiatric history before the index birth among those children whose biological father were identifiable. As parental psychiatric problem during childhood is a risk factor for child maltreatment^30^, we restricted analyses to offspring with no maternal psychiatric disorders records after birth, aiming to examine whether the observed association was purely through this hypothesized postnatal pathway. We also hypothesized another postnatal pathway: given the high heritability of psychiatric disorders and their frequent comorbidity with CVD, the observed association may be purely due to a higher risk of psychiatric disorders among offspring. Thus, we repeated our main model restricting to offspring without psychiatric diagnoses. To investigate potential moderating role of genetic predisposition to CVD,^31^ we performed stratified analyses based on maternal family history of CVD. To explore whether the association between maternal psychiatric disorders and the risk of overall CVD varied by offspring age, we conducted stratified analyses by splitting the follow-up at the attained ages of 10, 20, 30, and 40 years. To address the imbalance in certain covariates, particularly birth year, between the exposed and unexposed offspring, we created a matched sub-cohort using 1:1 nearest neighbor propensity score matching (PSM).^32^ The PSM method is described in detail in eTable 3. We then applied the same Cox model as in the full cohort to estimate the association between maternal psychiatric disorders and the risk of overall and subtype-specific CVD in the matched sub-cohort. Additional adjustment for maternal smoking and BMI during the index pregnancy was conducted among those with complete covariate data. Considering potential overadjustment for pregnancy complications in our full model, we removed hypertensive disorders and diabetes before or during the index pregnancy in additional analysis for comparison. To examine potential sex-specific associations or birth cohort effects, we further stratified the analyses by offspring sex and birth year (before 2001 versus 2001 or later).

## Results

Among the 4,171,005 singletons born alive between 1973 and 2014, 208,680 (5.0%) were born to mothers with a record of psychiatric disorder. Compared to the unexposed offspring, those born to mothers with psychiatric disorders were more likely to be born in later years of the study period, be born preterm, have congenital heart disease, and have a father with psychiatric disorder. Their mothers were more likely to be born in Sweden, be older and single at the time of childbirth, have lower educational attainment and parity of three or higher. These mothers were also more likely to smoke, have obesity during early pregnancy, and have hypertensive disorders, diabetes, and a family history of cardiovascular disease prior to childbirth (Table 1).

**Table 1.** Baseline characteristics of study participants.

|  | Exposure to maternal psychiatric disorders |  |
| --- | --- | --- |
|  | No<br>(N=3,962,325)<br>N (%) | Yes<br>(N=208,680)<br>N (%) |
| **Offspring birth year** |  |  |
| 1973-1978 | 588,884 (14.9) | 6023 (2.9) |
| 1979-1984 | 535,073 (13.5) | 12,237 (5.9) |
| 1985-1990 | 617,025 (15.6) | 16,776 (8.0) |
| 1991-1996 | 626,665 (15.8) | 18,734 (9.0) |
| 1997-2002 | 492,412 (12.4) | 19,611 (9.4) |
| 2003-2008 | 545,766 (13.8) | 45,551 (21.8) |
| 2009-2014 | 556,500 (14.0) | 89,748 (43.0) |
| **Offspring sex** |  |  |
| Male | 2,037,213 (51.4) | 107,267 (51.4) |
| Female | 1,925,112 (48.6) | 101,413 (48.6) |
| **Offspring preterm birth** |  |  |
| No | 3,762,144 (95.0) | 193,832 (92.9) |
| Yes | 191,228 (4.8) | 14,472 (6.9) |
| Unknown | 8953 (0.2) | 376 (0.2) |
| **Small for gestational age in offspring** |  |  |
| No | 697,878 (17.6) | 38,906 (18.6) |
| Yes | 3,246,850 (81.9) | 168,932 (81.0) |
| Unknown | 17,597 (0.4) | 842 (0.4) |
| **Congenital heart disease in offspring** |  |  |
| No | 3,904,730 (98.6) | 203,914 (97.7) |
| Yes | 57,595 (1.4) | 4766 (2.3) |
| **Maternal country of origin** |  |  |
| Sweden | 3,316,582 (83.7) | 178,392 (85.5) |
| Other countries | 643,911 (16.3) | 30,105 (14.4) |
| Unknown | 1832 (0.1) | 183 (0.1) |
| **Maternal age at the time of the study participant's birth (years)** |  |  |
| ≤19 | 119,533 (3.0) | 7375 (3.5) |
| 20-24 | 794,691 (20.1) | 40,346 (19.3) |
| 25-29 | 1,381,601 (34.9) | 61,958 (29.7) |
| 30-34 | 1,579,568 (39.9) | 90,704 (43.5) |
| ≥35 | 86,932 (2.2) | 8297 (4.0) |
| **Maternal education at the time of the study participant's birth** |  |  |
| Primary and lower secondary | 651,593 (16.4) | 53,086 (25.4) |
| Upper secondary | 1,859,583 (46.9) | 92,166 (44.2) |
| Bachelor or higher | 1,341,112 (33.9) | 59,700 (28.6) |
| Unknown | 110,037 (2.8) | 3728 (1.8) |
| **Maternal marital status at the time of the study participant's birth** |  |  |
| Not married/not in registered partnership | 1,897,373 (47.9) | 66,662 (31.9) |
| Married/registered partnership | 1,826,640 (46.1) | 133,145 (63.8) |
| Unknown | 238,312 (6.0) | 8873 (4.3) |
| **Maternal parity at the time of the study participant's birth** |  |  |
| 1 | 1,690,933 (42.7) | 89,465 (42.9) |
| 2 | 1,456,086 (36.8) | 68,076 (32.6) |
| ≥3 | 815,306 (20.6) | 51,139 (24.5) |
| **Maternal smoking in the first trimester of pregnancy <sup>a</sup>** |  |  |
| No | 2,449,857 (61.8) | 140,791 (67.5) |
| Yes | 447,143 (11.3) | 45,270 (21.7) |
| Unknown | 1,065,325 (26.9) | 22,619 (10.8) |
| **Maternal body-mass index in the first trimester of pregnancy (kg/m<sup>2</sup>) <sup>a</sup>** |  |  |
| <18.5 | 91,109 (2.3) | 6432 (3.1) |
| 18.5-24.9 | 1,584,762 (40.0) | 97,687 (46.8) |
| 25.0-29.9 | 502,366 (12.7) | 39,759 (19.1) |
| ≥30.0 | 199,101 (5.0) | 22,108 (10.6) |
| Unknown | 1,584,987 (40.0) | 42,694 (20.5) |
| **Maternal hypertensive disease before or during the index pregnancy** |  |  |
| No | 3,836,536 (96.8) | 199,962 (95.8) |
| Yes | 125,789 (3.2) | 8718 (4.2) |
| **Maternal diabetes before or during the index pregnancy** |  |  |
| No | 3,924,987 (99.1) | 204,534 (98.0) |
| Yes | 37,338 (0.9) | 4146 (2.0) |
| **Mother's family history of CVD before the birth of the study participant** |  |  |
| No | 3,555,541 (89.7) | 152,447 (73.1) |
| Yes | 406,784 (10.3) | 56,233 (26.9) |
| **Father's history of psychiatric disorders before the birth of the study participant** |  |  |
| No | 3,183,148 (80.3) | 145,048 (69.5) |
| Yes | 110,231 (2.8) | 26,040 (12.5) |
| Unknown | 668,946 (16.9) | 37,592 (18.0) |

Abbreviations: N, number; CVD, cardiovascular disease.
<sup>a</sup> Information on maternal smoking was collected since 1982; data on body-mass index was collected during 1982-1989 and 1992-2014.

Over the up to 51 years of follow-up (median, 27.6 years), 307,596 offspring (7.4%) were diagnosed with CVD. Throughout the follow-up period, offspring of mothers with psychiatric disorders had consistently higher overall CVD incidence rates than unexposed offspring (HR 1.19 [95% CI 1.17-1.21]; Figure 1, eFigure 1). In the cousin comparison cohort of 1,577,113 offspring, the association was consistently observed (HR 1.08 [1.03-1.13]), though the effect size was slightly attenuated. Sensitivity analyses in the full cohort showed a robust association when additionally adjusting for paternal psychiatric disorders, restricting to offspring without maternal diagnosis of psychiatric disorders after birth, and restricting to offspring without own diagnosis of psychiatric disorders (eTable 4). The association remained consistent across different follow-up intervals (<10 years, 10-19 years, 20-29 years, 30-39 years, and ≥40 years).

**Figure 1.**
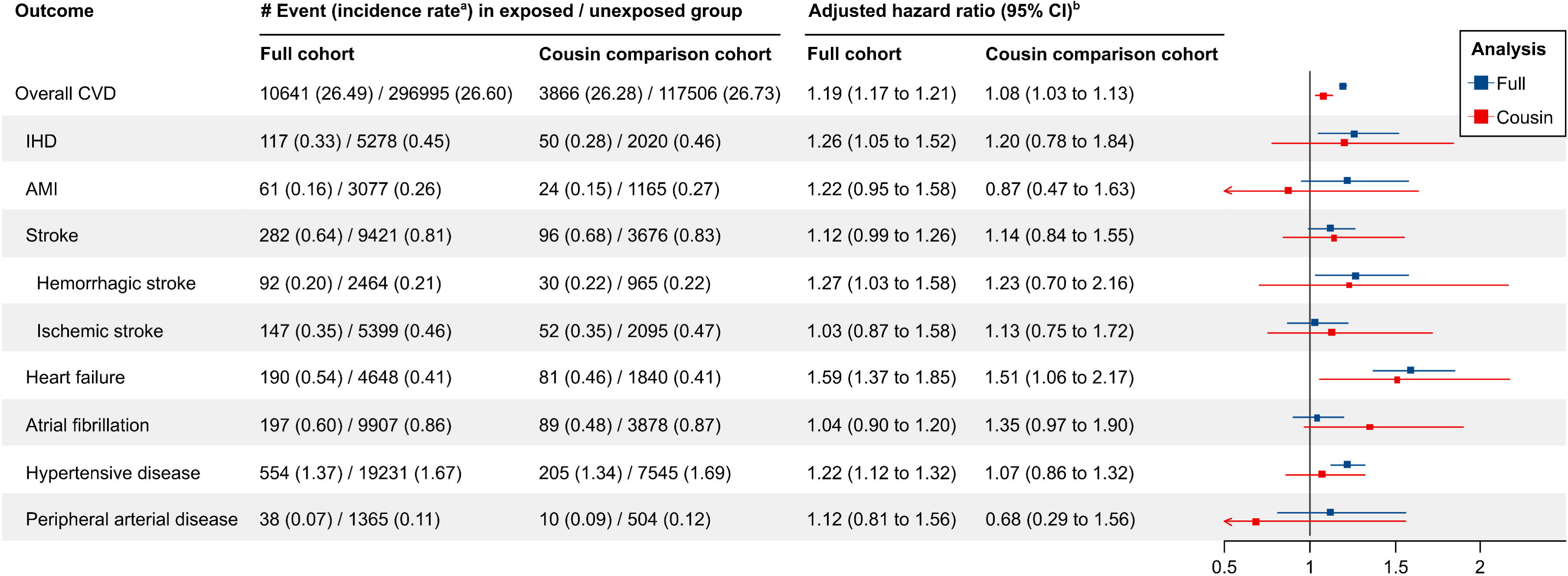
Association of maternal psychiatric disorders with overall and specific cardiovascular diseases in offspring in the full (N=4,171,005) and cousin comparison cohort (N=1,577,113). ^a^ Incidence rates are expressed as events per 100,00 person-years. ^b^ Models was adjusted for maternal country of origin, parity, maternal age at delivery, maternal education, maternal marital status, mother’s family history of cardiovascular diseases prior to the index birth, maternal hypertensive disorders and diabetes before or during the index pregnancy, offspring’s sex, and birth year. Notably, family history was inherently controlled for by the cousin-comparison design, thus was not adjusted for in corresponding models. Abbreviations: CI, confidence interval; CVD, cardiovascular disease; IHD, ischemic heart disease; AMI, acute myocardial infarction.

Among the studied CVD subtypes, the incidence rates of stroke, heart failure, and hypertensive disease in the exposed group began exceeding those in the unexposed group after around 20 years of follow-up. The elevated incidence rates of ischemic heart disease, acute myocardial infarction, and peripheral artery disease in the exposed group became apparent after about 35 years. The incidence rate of atrial fibrillation remained similar between the two groups across the follow-up period (eFigure 2-8). The most prominent association was observed between maternal psychiatric disorders and heart failure (HR 1.59 [1.37-1.85]; Figure 1; eTable 5), followed by ischemic heart disease, acute myocardial infarction, stroke (with stronger association for hemorrhagic than for ischemic stroke), and hypertensive disease. Notably, the association with heart failure was robust in the cousin-matched analysis (HR 1.51 [1.06-2.17]; Figure 1; eTable 6). The relative risk estimates for ischemic heart disease and hemorrhagic stroke were also similar to that of the full cohort.

Offspring CVD risk was increased across nearly all categories of maternal psychiatric disorders (Figure 2). The strongest associations were observed for maternal somatoform disorders (HR 1.35 [1.23-1.48]; eTable 7), which persisted in the cousin-matched analysis (HR 1.55 [1.23-1.97]; Figure 1; eTable 8). Maternal neurotic disorders showed consistent associations with offspring CVD across both the full cohort (HR 1.25, [1.21-1.28]) and cousin-matched analyses (HR 1.11, [1.03-1.19]), though most other associations were attenuated after adjustment for shared familial confounding in the cousin-comparison models.

**Figure 2.**
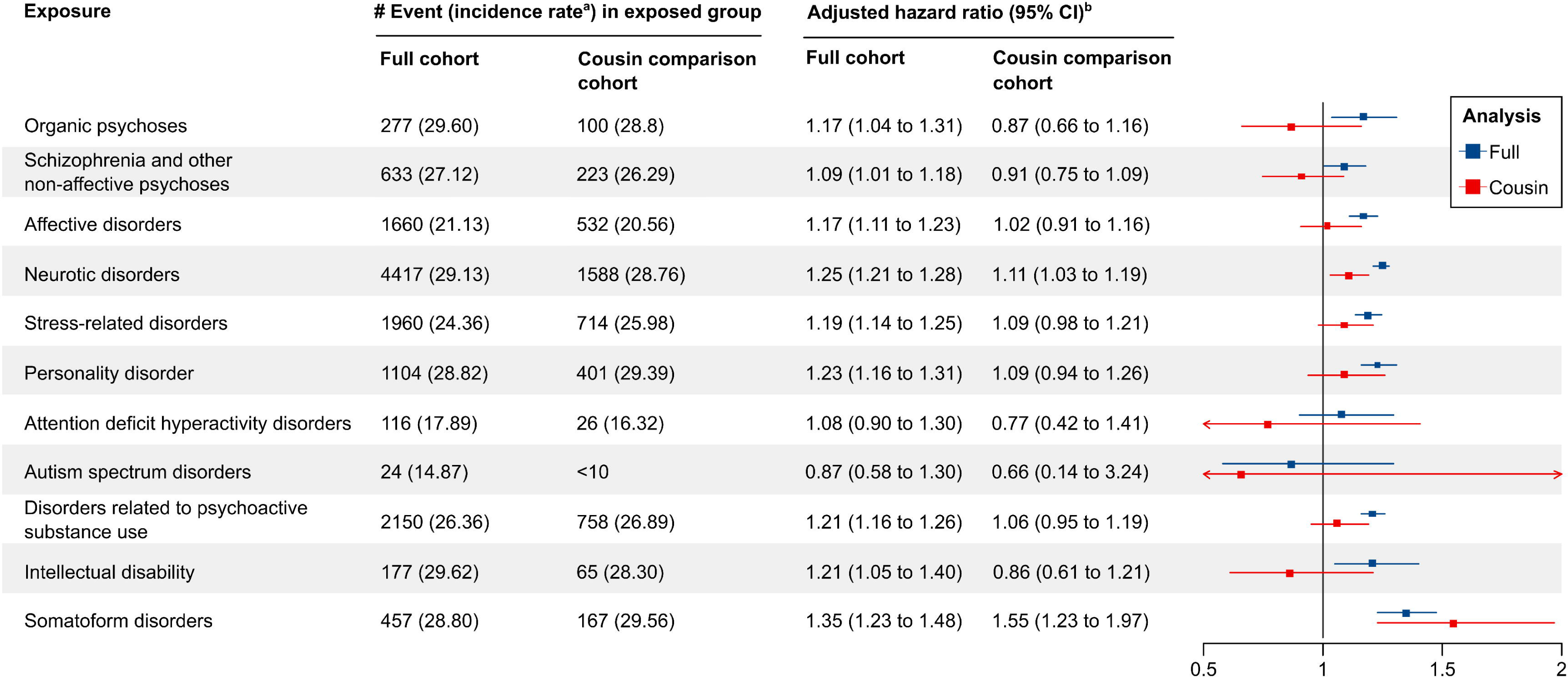
Association of maternal psychiatric disorder subtypes with overall cardiovascular diseases in offspring in the full (N=4,171,005) and cousin comparison cohort (N=1,577,113). ^a^ Incidence rates are expressed as events per 100,00 person-years. ^b^ Models was adjusted for maternal country of origin, parity, maternal age at delivery, maternal education, maternal marital status, mother’s family history of cardiovascular diseases prior to the index birth, maternal hypertensive disorders and diabetes before or during the index pregnancy, offspring’s sex, and birth year. Notably, family history was inherently controlled for by the cousin-comparison design, thus was not adjusted for in corresponding models. Abbreviations: CI, confidence interval.

The mediation analysis indicated that congenital heart disease accounted for a substantial proportion of the association between maternal psychiatric disorders and the risk of CVD (proportion mediated, 9.5%), particularly in case of stroke (proportion mediated, 34.3%), ischemic heart disease, and heart failure (both proportion mediated ∼15%; Table 2). In contrast, preterm birth and small for gestational age contributed minimally to these associations (proportion mediated < 3% for the composite CVD outcome).

**Table 2.**
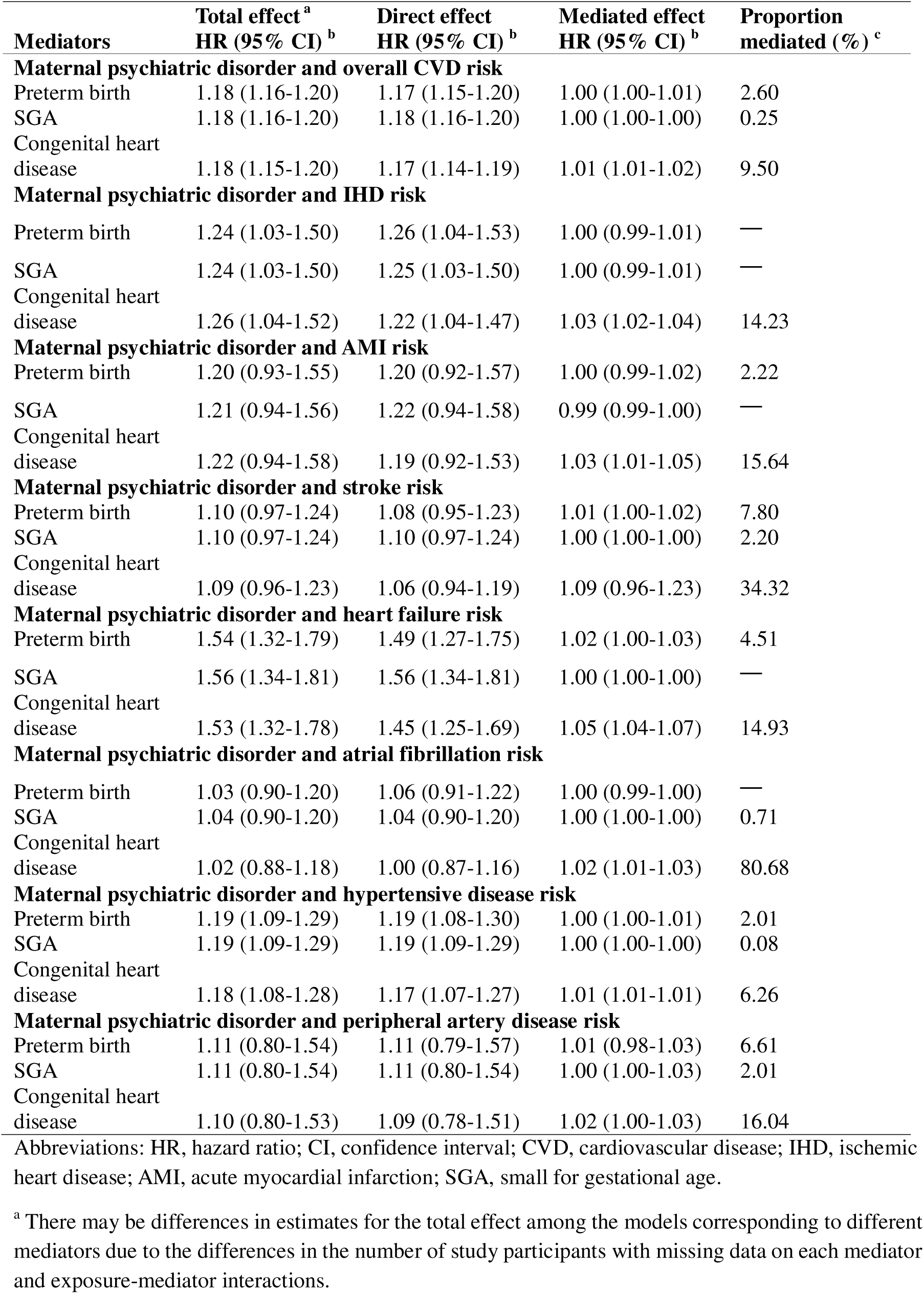
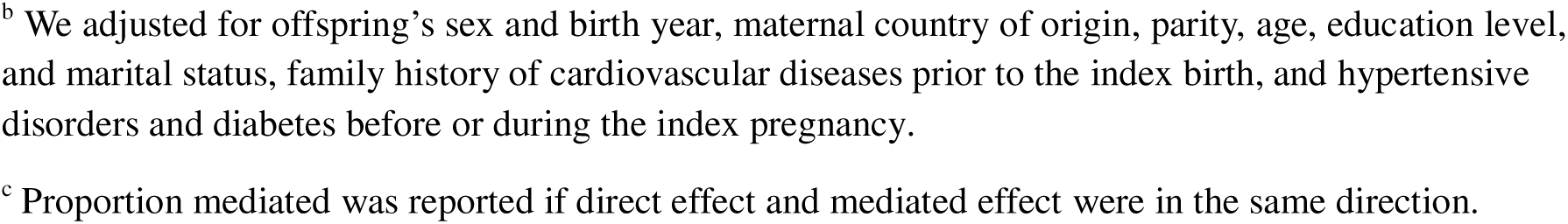
Mediating roles of preterm birth, small for gestational age, and congenital heart disease in the association between maternal psychiatric disorder and risk of overall cardiovascular disease.

| **Mediators** | **Total effect <sup>a</sup>**<br>**HR (95% CI) <sup>b</sup>** | **Direct effect**<br>**HR (95% CI) <sup>b</sup>** | **Mediated effect**<br>**HR (95% CI) <sup>b</sup>** | **Proportion**<br>**mediated (%) <sup>c</sup>** |
| --- | --- | --- | --- | --- |
| **Maternal psychiatric disorder and overall CVD risk** |  |  |  |  |
| Preterm birth | 1.18 (1.16-1.20) | 1.17 (1.15-1.20) | 1.00 (1.00-1.01) | 2.60 |
| SGA | 1.18 (1.16-1.20) | 1.18 (1.16-1.20) | 1.00 (1.00-1.00) | 0.25 |
| Congenital heart disease | 1.18 (1.15-1.20) | 1.17 (1.14-1.19) | 1.01 (1.01-1.02) | 9.50 |
| **Maternal psychiatric disorder and IHD risk** |  |  |  |  |
| Preterm birth | 1.24 (1.03-1.50) | 1.26 (1.04-1.53) | 1.00 (0.99-1.01) | — |
| SGA | 1.24 (1.03-1.50) | 1.25 (1.03-1.50) | 1.00 (0.99-1.01) | — |
| Congenital heart disease | 1.26 (1.04-1.52) | 1.22 (1.04-1.47) | 1.03 (1.02-1.04) | 14.23 |
| **Maternal psychiatric disorder and AMI risk** |  |  |  |  |
| Preterm birth | 1.20 (0.93-1.55) | 1.20 (0.92-1.57) | 1.00 (0.99-1.02) | 2.22 |
| SGA | 1.21 (0.94-1.56) | 1.22 (0.94-1.58) | 0.99 (0.99-1.00) | — |
| Congenital heart disease | 1.22 (0.94-1.58) | 1.19 (0.92-1.53) | 1.03 (1.01-1.05) | 15.64 |
| **Maternal psychiatric disorder and stroke risk** |  |  |  |  |
| Preterm birth | 1.10 (0.97-1.24) | 1.08 (0.95-1.23) | 1.01 (1.00-1.02) | 7.80 |
| SGA | 1.10 (0.97-1.24) | 1.10 (0.97-1.24) | 1.00 (1.00-1.00) | 2.20 |
| Congenital heart disease | 1.09 (0.96-1.23) | 1.06 (0.94-1.19) | 1.09 (0.96-1.23) | 34.32 |
| **Maternal psychiatric disorder and heart failure risk** |  |  |  |  |
| Preterm birth | 1.54 (1.32-1.79) | 1.49 (1.27-1.75) | 1.02 (1.00-1.03) | 4.51 |
| SGA | 1.56 (1.34-1.81) | 1.56 (1.34-1.81) | 1.00 (1.00-1.00) | — |
| Congenital heart disease | 1.53 (1.32-1.78) | 1.45 (1.25-1.69) | 1.05 (1.04-1.07) | 14.93 |
| **Maternal psychiatric disorder and atrial fibrillation risk** |  |  |  |  |
| Preterm birth | 1.03 (0.90-1.20) | 1.06 (0.91-1.22) | 1.00 (0.99-1.00) | — |
| SGA | 1.04 (0.90-1.20) | 1.04 (0.90-1.20) | 1.00 (1.00-1.00) | 0.71 |
| Congenital heart disease | 1.02 (0.88-1.18) | 1.00 (0.87-1.16) | 1.02 (1.01-1.03) | 80.68 |
| **Maternal psychiatric disorder and hypertensive disease risk** |  |  |  |  |
| Preterm birth | 1.19 (1.09-1.29) | 1.19 (1.08-1.30) | 1.00 (1.00-1.01) | 2.01 |
| SGA | 1.19 (1.09-1.29) | 1.19 (1.09-1.29) | 1.00 (1.00-1.00) | 0.08 |
| Congenital heart disease | 1.18 (1.08-1.28) | 1.17 (1.07-1.27) | 1.01 (1.01-1.01) | 6.26 |
| **Maternal psychiatric disorder and peripheral artery disease risk** |  |  |  |  |
| Preterm birth | 1.11 (0.80-1.54) | 1.11 (0.79-1.57) | 1.01 (0.98-1.03) | 6.61 |
| SGA | 1.11 (0.80-1.54) | 1.11 (0.80-1.54) | 1.00 (1.00-1.03) | 2.01 |
| Congenital heart disease | 1.10 (0.80-1.53) | 1.09 (0.78-1.51) | 1.02 (1.00-1.03) | 16.04 |

Abbreviations: HR, hazard ratio; CI, confidence interval; CVD, cardiovascular disease; IHD, ischemic heart disease; AMI, acute myocardial infarction; SGA, small for gestational age.
<sup>a</sup> There may be differences in estimates for the total effect among the models corresponding to different mediators due to the differences in the number of study participants with missing data on each mediator and exposure-mediator interactions.
<sup>b</sup> We adjusted for offspring's sex and birth year, maternal country of origin, parity, age, education level, and marital status, family history of cardiovascular diseases prior to the index birth, and hypertensive disorders and diabetes before or during the index pregnancy.
<sup>c</sup> Proportion mediated was reported if direct effect and mediated effect were in the same direction.

We did not find find evidence of effect modification by maternal family history of CVD before birth, offspring sex and birth year (eTable 4). Sensitivity analyses yielded largely consistent findings with those in the main analyses, i.e. when additionally adjusting for maternal smoking and pre-pregnancy body-mass index (eTable 4), removing maternal pregnancy complications as covariates (eTable 5 and 7), and alternatively utilizing PSM to balance baseline characteristics (eTable 9).

## Discussion

In this nationwide population-based cohort study involving over 4.1 million individuals, maternal history of psychiatric disorders was associated with a moderately increased CVD risk in offspring from childhood up to middle-age. The association was consistent across most of the studied maternal psychiatric disorder subtypes and offspring CVD outcomes. In the cousin-comparison analysis, the estimates were attenuated but remained elevated. Congenital heart disease explained 9.5% of the observed association, while the mediating effect of adverse pregnancy outcomes appeared to be minimal.

To our knowledge, no previous study has systematically investigated the association between maternal psychiatric disorders and the risk of CVD in offspring. Our findings are in line with those of two earlier Nordic register-based studies showing that maternal bereavement, an objective source of severe stress, shortly before or during pregnancy, was associated with increased risks of overall CVD and of heart failure in offspring during childhood and early adulthood.^33^ Results from the present study are in part consistent with the findings of the few earlier studies in this area analyzing the association of maternal depressive symptoms and stress during pregnancy with cardiometabolic profile in offspring.^16,18,34^ Several of these studies have shown positive associations between maternal psychopathology during pregnancy and childhood overweight.^17,18^ Meanwhile, evidence for other cardiometabolic outcomes, e.g. blood pressure and central adiposity in childhood, has been either inconsistent or limited by insufficient sample size.^35–38^ Our study extends these earlier lines of investigations by analyzing in a large cohort a broad spectrum of maternal psychiatric disorders and of offspring CVD during an up to five decades follow-up.

Regarding potential mechanistic pathways, maternal psychiatric disorders have been shown to be associated with congenital malformations in offspring^16^, which in turn predispose the offspring to an increased risk of CVD.^13^ In this study, congenital heart disease emerged as a mediator of the studied association. While the precise mechanisms linking maternal psychiatric disorders to congenital heart disease remain to be fully elucidated, we observed a consistent association among the cousin-matched cohort, suggesting a pathway potentially independent of genetic factors. In addition to the recognized dysregulation of the hypothalamic-pituitary-adrenal axis,^6,7,39,40^ adverse health behaviors during pregnancy associated with psychiatry disorder may also play a role.^10,41^ Preventive strategies may include enhanced preconception counseling, optimization of modifiable risk factors such as folate supplementation, and adherence to recommended prenatal care protocols.^41^ Despite higher rates of adverse pregnancy outcomes among women with psychiatric disorders, small for gestational age and preterm birth played a limited role in explaining the increased offspring CVD risk.

The main strength of this study is the use of nationwide, population-based registers with almost complete follow-up from birth to mid-adulthood. The large sample size permitted a comprehensive evaluation of specific types of CVD and a cousin-comparison design to account for familial genetic and early-life environmental factors. Furthermore, the high validity of psychiatric and CVD diagnoses within the registry has been well-documented.^22^ Several limitations should also be noted. First, while various hypotheses regarding the underlying mechanisms were explored, we did not have data to investigate further potential explanations, such as medication use during pregnancy and postnatal factors.^42–44^ Second, despite the large cohort, the sample size was not sufficient to fully examine type-specific associations between certain maternal psychiatric disorders and specific CVD outcomes. Third, exposure misclassification was likely and could manifest in two distinct ways. One potential source involves psychiatric disorders that resolved before pregnancy, such as mild depression, without subsequent relapse during gestation. The second source of potential misclassification stems from our reliance on inpatient and specialized outpatient care records for identifying psychiatric disorders. This approach inadvertently classified women with mild to moderate symptoms who exclusively sought primary care treatment into the unexposed group and likely lead to an underestimation of the association.

In summary, maternal history of psychiatric disorders was associated with a moderately increased CVD risk in offspring from childhood through early adulthood. Congenital heart disease accounted for approximately 10% of this observed association, while adverse birth outcomes played relatively minor roles.

## Acknowledgment

Ethical approval:

The study was approved by the Research Ethics Committee at Karolinska Institutet in Stockholm (No. 2016/288-31/1, 2021-03315, 2025-04808-02). The boards do not request informed consent for register-based studies.

## Transparency statement

The lead author affirms that the manuscript is an honest, accurate, and transparent account of the study being reported. No important aspects of the study have been omitted.

## Author Contributions

FY and KL had full access to all the data in the study and take responsibility for the integrity of the data. JL, KL, FY and TR conceived and designed the study. FY performed the data management and the statistical analyses. KL provided administrative, technical, and material support. JL and KL supervised the study. All authors contributed to analyses or interpretation of the results. FY and TR drafted the manuscript, and all authors critically revised it for important intellectual content. The corresponding author attests that all listed authors meet authorship criteria and that no others meeting the criteria have been omitted.

## Data availability

We are not allowed to share data from the Swedish registers due to strict regulation and ethical considerations.

## References

1. GBD 2021 Diseases and Injuries Collaborators,. Global incidence, prevalence, years lived with disability (YLDs), disability-adjusted life-years (DALYs), and healthy life expectancy (HALE) for 371 diseases and injuries in 204 countries and territories and 811 subnational locations, 1990–2021: a systematic analysis for the Global Burden of Disease Study 2021. The Lancet. 2024;403(10440):2133–2161. doi:10.1016/S0140-6736(24)00757-8

2. Sun J, Qiao Y, Zhao M, Magnussen CG, Xi B. Global, regional, and national burden of cardiovascular diseases in youths and young adults aged 15–39 years in 204 countries/territories, 1990–2019: a systematic analysis of Global Burden of Disease Study 2019. BMC Med. 2023;21(1):222. doi:10.1186/s12916-023-02925-4

3. Yu Y, Arah OA, Liew Z, et al. Maternal diabetes during pregnancy and early onset of cardiovascular disease in offspring: population based cohort study with 40 years of follow-up. BMJ. 2019;367:l6398. doi:10.1136/bmj.l6398

4. Barker DJ, Bagby SP, Hanson MA. Mechanisms of Disease: in utero programming in the pathogenesis of hypertension. Nat Rev Nephrol. 2006;2(12):700–707. doi:10.1038/ncpneph0344

5. Solmi M, Radua J, Olivola M, et al. Age at onset of mental disorders worldwide: large-scale meta-analysis of 192 epidemiological studies. Mol Psychiatry. 2022;27(1):281–295. doi:10.1038/s41380-021-01161-7

6. Christian LM, Franco A, Glaser R, Iams JD. Depressive symptoms are associated with elevated serum proinflammatory cytokines among pregnant women. Brain, Behavior, and Immunity. 2009;23(6):750–754. doi:10.1016/j.bbi.2009.02.012

7. Liu MY, Wei LL, Zhu XH, et al. Prenatal stress modulates HPA axis homeostasis of offspring through dentate TERT independently of glucocorticoids receptor. Mol Psychiatry. 2023;28(3):1383–1395. doi:10.1038/s41380-022-01898-9

8. Moisiadis VG, Matthews SG. Glucocorticoids and fetal programming part 1: outcomes. Nat Rev Endocrinol. 2014;10(7):391–402. doi:10.1038/nrendo.2014.73

9. Wang S, Mei Z, Chen J, et al. Maternal Immune Activation: Implications for Congenital Heart Defects. Clinic Rev Allerg Immunol. 2025;68(1):36. doi:10.1007/s12016-025-09049-y

10. Míguez MC, Queiro Y, Posse CM, Val A. The Connection Between Stress and Women’s Smoking During the Perinatal Period: A Systematic Review. Brain Sciences. 2024;15(1):13. doi:10.3390/brainsci15010013

11. Sixtus RP, Dyson RM, Gray CL. Impact of prematurity on lifelong cardiovascular health: structural and functional considerations. npj Cardiovasc Health. 2024;1(1):2. doi:10.1038/s44325-024-00002-0

12. Crispi F, Miranda J, Gratacós E. Long-term cardiovascular consequences of fetal growth restriction: biology, clinical implications, and opportunities for prevention of adult disease. Am J Obstet Gynecol. 2018;218(2S):S869–S879. doi:10.1016/j.ajog.2017.12.012

13. Wang T, Chen L, Yang T, et al. Congenital Heart Disease and Risk of Cardiovascular Disease: A Meta-Analysis of Cohort Studies. J Am Heart Assoc. 2019;8(10):e012030. doi:10.1161/JAHA.119.012030

14. Ghimire U, Papabathini SS, Kawuki J, Obore N, Musa TH. Depression during pregnancy and the risk of low birth weight, preterm birth and intrauterine growth restriction- an updated meta-analysis. Early Hum Dev. 2021;152:105243. doi:10.1016/j.earlhumdev.2020.105243

15. Voit FAC, Kajantie E, Lemola S, Räikkönen K, Wolke D, Schnitzlein DD. Maternal mental health and adverse birth outcomes. PLoS One. 2022;17(8):e0272210. doi:10.1371/journal.pone.0272210

16. Gu J, Guan HB. Maternal psychological stress during pregnancy and risk of congenital heart disease in offspring: A systematic review and meta-analysis. Journal of Affective Disorders. 2021;291:32–38. doi:10.1016/j.jad.2021.05.002

17. Benton PM, Skouteris H, Hayden M. Does maternal psychopathology increase the risk of pre-schooler obesity? A systematic review. Appetite. 2015;87:259–282. doi:10.1016/j.appet.2014.12.227

18. Eberle C, Fasig T, Brüseke F, Stichling S. Impact of maternal prenatal stress by glucocorticoids on metabolic and cardiovascular outcomes in their offspring: A systematic scoping review. Xie L, ed. PLoS ONE. 2021;16(1):e0245386. doi:10.1371/journal.pone.0245386

19. Ludvigsson JF, Almqvist C, Bonamy AKE, et al. Registers of the Swedish total population and their use in medical research. Eur J Epidemiol. 2016;31(2):125–136. doi:10.1007/s10654-016-0117-y

20. Axelsson O. The Swedish medical birth register. Acta Obstet Gynecol Scand. 2003;82(6):491–492. doi:10.1034/j.1600-0412.2003.00172.x

21. Socialstyrelsen. National Patient Register. Accessed June 3, 2025. https://www.socialstyrelsen.se/en/statistics-and-data/registers/national-patient-register/

22. Everhov ÅH, Frisell T, Osooli M, et al. Diagnostic accuracy in the Swedish national patient register: a review including diagnoses in the outpatient register. Eur J Epidemiol. 2025;40(3):359–369. doi:10.1007/s10654-025-01221-0

23. Frederiksen LE, Erdmann F, Mader L, et al. Psychiatric disorders in childhood cancer survivors in Denmark, Finland, and Sweden: a register-based cohort study from the SALiCCS research programme. Lancet Psychiatry. 2022;9(1):35–45. doi:10.1016/S2215-0366(21)00387-4

24. Brooke HL, Talbäck M, Hörnblad J, et al. The Swedish cause of death register. Eur J Epidemiol. 2017;32(9):765–773. doi:10.1007/s10654-017-0316-1

25. Shen Q, Mikkelsen DH, Luitva LB, et al. Psychiatric disorders and subsequent risk of cardiovascular disease: a longitudinal matched cohort study across three countries. eClinicalMedicine. 2023;61:102063. doi:10.1016/j.eclinm.2023.102063

26. Lindekilde N, Scheuer SH, Diaz LJ, et al. Risk of Developing Type 2 Diabetes in Individuals With a Psychiatric Disorder: A Nationwide Register-Based Cohort Study. Diabetes Care. 2022;45(3):724–733. doi:10.2337/dc21-1864

27. Huang C, Li J, Qin G, et al. Maternal hypertensive disorder of pregnancy and offspring early-onset cardiovascular disease in childhood, adolescence, and young adulthood: A national population-based cohort study. Bassat Q, ed. PLoS Med. 2021;18(9):e1003805. doi:10.1371/journal.pmed.1003805

28. Marsál K, Persson PH, Larsen T, Lilja H, Selbing A, Sultan B. Intrauterine growth curves based on ultrasonically estimated foetal weights. Acta Paediatr. 1996;85(7):843–848. doi:10.1111/j.1651-2227.1996.tb14164.x

29. Valeri L, Vanderweele TJ. Mediation analysis allowing for exposure-mediator interactions and causal interpretation: theoretical assumptions and implementation with SAS and SPSS macros. Psychol Methods. 2013;18(2):137–150. doi:10.1037/a0031034

30. Nevriana A, Kosidou K, Hope H, et al. Parental Mental Illness and the Likelihood of Child Out-of-Home Care: A Cohort Study. Pediatrics. 2024;153(3):e2023061531. doi:10.1542/peds.2023-061531

31. Hossin MZ, Kazamia K, Faxén J, et al. Pre-existing maternal cardiovascular disease and the risk of offspring cardiovascular disease from infancy to early adulthood. European Heart Journal. 2024;45(38):4111–4123. doi:10.1093/eurheartj/ehae547

32. Ho DE, Imai K, King G, Stuart EA. **MatchIt**L: Nonparametric Preprocessing for Parametric Causal Inference. J Stat Soft. 2011;42(8). doi:10.18637/jss.v042.i08

33. Plana-Ripoll O, Liu X, Momen NC, Parner E, Olsen J, Li J. Prenatal exposure to maternal stress following bereavement and cardiovascular disease: A nationwide population-based and sibling-matched cohort study. Eur J Prev Cardiolog. 2016;23(10):1018–1028. doi:10.1177/2047487315585294

34. Doom JR, Deer LK, Dabelea D, et al. Biological and behavioral pathways from prenatal depression to offspring cardiometabolic risk: Testing the developmental origins of health and disease hypothesis. Developmental Psychology. 2024;60(9):1620–1638. doi:10.1037/dev0001704

35. Ertel KA, Koenen KC, Rich-Edwards JW, Gillman MW. Antenatal and postpartum depressive symptoms are differentially associated with early childhood weight and adiposity. Paediatric and Perinatal Epidemiology. 2010;24(2):179–189. doi:10.1111/j.1365-3016.2010.01098.x

36. Van Dijk AE, Van Eijsden M, Stronks K, Gemke RJBJ, Vrijkotte TGM. The Association between Prenatal Psychosocial Stress and Blood Pressure in the Child at Age 5–7 Years. PLoS ONE. 2012;7(8):e43548. doi:10.1371/journal.pone.0043548

37. Van Dijk AE, Dawe K, Deanfield J, et al. The association of maternal prenatal psychosocial stress with vascular function in the child at age 10–11 years: findings from the Avon longitudinal study of parents and children. Eur J Prev Cardiolog. 2014;21(9):1097–1108. doi:10.1177/2047487313486039

38. Silva CCV, Vehmeijer FOL, El Marroun H, Felix JF, Jaddoe VWV, Santos S. Maternal psychological distress during pregnancy and childhood cardio-metabolic risk factors. *Nutrition*, Metabolism and Cardiovascular Diseases. 2019;29(6):572–579. doi:10.1016/j.numecd.2019.02.008

39. Irwin JL, Meyering AL, Peterson G, et al. Maternal prenatal cortisol programs the infant hypothalamic–pituitary–adrenal axis. Psychoneuroendocrinology. 2021;125:105106. doi:10.1016/j.psyneuen.2020.105106

40. O’Connor TG, Tang W, Gilchrist MA, Moynihan JA, Pressman EK, Blackmore ER. Diurnal cortisol patterns and psychiatric symptoms in pregnancy: Short-term longitudinal study. Biological Psychology. 2014;96:35–41. doi:10.1016/j.biopsycho.2013.11.002

41. Chowdhury D, Elliott PA, Asaki SY, et al. Addressing Disparities in Pediatric Congenital Heart Disease: A Call for Equitable Health Care. JAHA. 2024;13(13):e032415. doi:10.1161/JAHA.123.032415

42. Nevriana A, Pierce M, Dalman C, et al. Association between maternal and paternal mental illness and risk of injuries in children and adolescents: nationwide register based cohort study in Sweden. BMJ. 2020;369:m853. doi:10.1136/bmj.m853

43. Shalev A, Merranko J, Goldstein T, et al. A Longitudinal Study of Family Functioning in Offspring of Parents Diagnosed With Bipolar Disorder. Journal of the American Academy of Child & Adolescent Psychiatry. 2019;58(10):961–970. doi:10.1016/j.jaac.2018.10.011

44. Heradstveit O, Haugland BSM, Nilsen SA, Bøe T, Sivertsen B, Hysing M. Parental Mental Illness as a Risk Factor for Adolescent Psychiatric Disorders: A Registry-Based Study of Specialized Child and Adolescent Health Services. Child & Youth Services. 2023;44(1):48–71. doi:10.1080/0145935X.2021.1997584

