## Supplementary Materials for "Association between maternal history of psychiatric disorders and the risk of cardiovascular disease in offspring up to early middle-age"

**Association of maternal psychiatric disorders before and during pregnancy with the risk of cardiovascular disease in offspring up to early middle-age**

**Supplementary Materials**

### eTable 1. Short description of the registers used in the study

| **Data source** | **Information retrieved from the register** | **Period covered by the register and our study** |
| --- | --- | --- |
| Total Population Register | Maternal country of origin, marital status, and index person’s migration | 1968-2014 |
| Multi-Generation Register | Family relationships for all mothers born since 1932 | 1961-2014 |
| Education Register | Maternal highest education | 1985-2014 |
| Medical Birth Register | Index person’s birth date, sex, gestational age, birth weight, singleton or multiple pregnancy, and maternal age at delivery, parity, height, weight and smoking status in early pregnancy, and complications during index pregnancy | 1973-2014 |
| Patient Register ^a^ | Date and diagnosis for inpatient and outpatient care | Inpatient care: 1964-2023 (its coverage became nationwide in 1987); specialised outpatient care: 2001-2023 |
| Cause of Death Register | Date and causes of death | 1952-2023 |

^a^ Before 1987, the coverage of psychiatric care in the Inpatient Register was higher than that of somatic care: in the year 1984-1985, 86% of all somatic care and 94% of all psychiatric inpatient care were reported to the Patient Register.

### eTable 2. International classification of diseases codes searched for to define the variables in our study

| **Disease or cause of death** | **ICD-8** | **ICD-9** | **ICD-10** |
| --- | --- | --- | --- |
| **Psychiatric disorders** | 290-315 | 290-319 | F00-F99 |
| **Subtypes of psychiatric disorders** | | | |
| Organic psychoses | 290-294, 309 | 290-294, 310 | F00-F09 |
| Schizophrenia and other non-affective psychoses | 295，297，298 | 295，297，298 | F20-F29 |
| Affective disorders | 296 | 296 | F30-F39 |
| Neuroses | 300 | 300 | F40-F42, F44, F48 |
| Stress-related disorders | 307 | 308-309 | F43 |
| Personality disorder | 301 | 301 | F60-F69 |
| Attention deficit hyperactivity disorders | null | 314 | F90 |
| Autism spectrum disorders | null | 299 | F84 |
| Disorders related to psychoactive substance use | 303, 304 | 303,304,305 | F10-F19 |
| Intellectual disability | 310-315 | 317-319 | F70-F79 |
| Somatoform disorders | 305 | 306 | F45 |
| **Cardiovascular disease** | 390-458 | 390-459 | I00-I99 |
| **Subtypes of cardiovascular disease** | | | |
| Ischemic heart disease | 410-414 | 410-414 | I20-I25 |
| Acute myocardial infarction | 410 | 410 | I21, I22 |
| Stroke | 430, 431, 433, 434, 436 | 430, 431, 433, 434, 436 | I60, I61, I63, I64 |
| Haemorrhagic stroke | 431 | 431 | I61 |
| Ischemic stroke | 433, 434 | 433, 434 | I63, I64 |
| Heart failure | 427.00, 427.10 | 428 | I11.0, I13.0, I13.2, I50 |
| Atrial fibrillation | 427.92 | 427D | I48 |
| Hypertensive disease | 400-404, 63701, 63703, 63704, 63709, 63799, 63710 | 401-405, 642A-642H, 642X | I10-I15, O10, O11, O13, O14, O15, O16 |
| Peripheral arterial disease | 440, 4438, 4439, 445 | 440, 443W, 443X | I70, I73.9 |
| **Congenital heart defects** | 746, 747 | 745, 746, 747 | Q20-Q28 |
| **Diabetes** | 250 | 250, 648A, 648W | E10-E14, O24 |

Abbreviation: ICD, International Statistical Classification of Diseases and Related Health Problems.

### eTable 3. Baseline characteristics of study participants in the propensity-score-matched sub-cohort

|  | **Maternal history of psychiatric disorders** | |
| --- | --- | --- |
|  | **No**  **(N=208,680)** | **Yes**  **(N=208,680)** |
|  | **N (%)** | **N (%)** |
| **Offspring birth year** | | |
| 1973-1978 | 6022 (2.9) | 6023 (2.9) |
| 1979-1984 | 12,239 (5.9) | 12,237 (5.9) |
| 1985-1990 | 16,782 (8.0) | 16,776 (8.0) |
| 1991-1996 | 18,761 (9.0) | 18,734 (9.0) |
| 1997-2002 | 19,704 (9.4) | 19,611 (9.4) |
| 2003-2008 | 45,696 (21.9) | 45,551 (21.8) |
| 2009-2014 | 89,476 (42.9) | 89,748 (43.0) |
| **Offspring sex** | | |
| Male | 107,320 (51.4) | 107,267 (51.4) |
| Female | 101,360 (48.6) | 101,413 (48.6) |
| **Offspring preterm birth** | | |
| No | 198,040 (94.9) | 193,832 (92.9) |
| Yes | 10,402 (5.0) | 14,472 (6.9) |
| Unknown | 238 (0.1) | 376 (0.2) |
| **Small for gestational age in offspring** | | |
| No | 37625 (18.0) | 38,906 (18.6) |
| Yes | 170426 (81.7) | 168,932 (81.0) |
| Unknown | 629 (0.3) | 842 (0.4) |
| **Congenital heart disease in offspring** | | |
| No | 204,825 (98.2) | 203,914 (97.7) |
| Yes | 3855 (1.8) | 4766 (2.3) |
| **Maternal country of origin** | | |
| Sweden | 178,494 (85.5) | 178,392 (85.5) |
| Other countries | 30,037 (14.4) | 30,105 (14.4) |
| Unknown | 149 (0.1) | 183 (0.1) |
| **Maternal age at the time of the study participant’s birth (years)** | | |
| ≤19 | 7322 (3.5) | 7375 (3.5) |
| 20-24 | 40,178 (19.3) | 40,346 (19.3) |
| 25-29 | 61,985 (29.7) | 61,958 (29.7) |
| 30-34 | 90,948 (43.6) | 90,704 (43.5) |
| ≥35 | 8247 (4.0) | 8297 (4.0) |
| **Maternal education at the time of the study participant’s birth** | | |
| Primary and lower secondary | 53,396 (25.6) | 53,086 (25.4) |
| Upper secondary | 92,237 (44.2) | 92,166 (44.2) |
| Bachelor or higher | 59,623 (28.6) | 59,700 (28.6) |
| Unknown | 3424 (1.6) | 3728 (1.8) |
| **Maternal marital status at the time of the study participant’s birth** | | |
| Not married/not in registered partnership | 66,632 (31.9) | 66,662 (31.9) |
| Married/registered partnership | 133,239 (63.8) | 133,145 (63.8) |
| Unknown | 8809 (4.2) | 8873 (4.3) |
| **Maternal parity** **at the time of the study participant’s birth** | | |
| 1 | 89,272 (42.8) | 89,465 (42.9) |
| 2 | 68,187 (32.7) | 68,076 (32.6) |
| ≥3 | 51,221 (24.5) | 51,139 (24.5) |
| **Maternal smoking in the first trimester of pregnancy ^a^** | | |
| No | 159,211 (76.3) | 140,791 (67.5) |
| Yes | 27,637 (13.2) | 45,270 (21.7) |
| Unknown | 21,832 (10.5) | 22,619 (10.8) |
| **Maternal body-mass index in the first trimester of pregnancy (kg/m^2^) ^a^** | | |
| <18.5 | 5160 (2.5) | 6432 (3.1) |
| 18.5-24.9 | 102357 (49.0) | 97,687 (46.8) |
| 25.0-29.9 | 40,394 (19.4) | 39,759 (19.1) |
| ≥30.0 | 20,569 (9.9) | 22,108 (10.6) |
| Unknown | 40,200 (19.3) | 42,694 (20.5) |
| **Maternal hypertensive disease before or during the index pregnancy** | | |
| No | 200,249 (96.0) | 199,962 (95.8) |
| Yes | 8431 (4.0) | 8718 (4.2) |
| **Maternal diabetes before or during the index pregnancy** | | |
| No | 204,763 (98.1) | 204,534 (98.0) |
| Yes | 3917 (1.9) | 4146 (2.0) |
| **Mother’s family history of cardiovascular disease before the index birth** | | |
| No | 152,527 (73.1) | 152,447 (73.1) |
| Yes | 56,153 (26.9) | 56,233 (26.9) |

Abbreviations: N, number.

We estimated the propensity score using logistic regression, modeling the probability of maternal psychiatric disorder as a function of the following covariates: offspring’s birth year and sex, maternal country of origin, parity, age, education level, marital status, family history of cardiovascular diseases prior to the index birth, and hypertensive disorders and diabetes before or during the index pregnancy. Covariate balance in the matched sample was assessed using standardized mean differences, all of which were below 0.1, indicating adequate balance.

### eTable 4. Additional analyses on the association between maternal psychiatric disorder and risk of overall cardiovascular diseases.

| **Group** | **No. of CVD cases** | **Incident CVD rate (10,000 person-years)** | **Crude HR**  **(95% CI)** | **Adjusted HR**  **(95% CI)^a^** |
| --- | --- | --- | --- | --- |
| **Additional adjustment for paternal psychiatric disorders before the index birth^b^** | | | | |
| Unexposed | 263,635 | 27.32 | 1.00 (ref) | 1.00 (ref) |
| Exposed | 8969 | 26.51 | 1.40 (1.37-1.43) | 1.17 (1.15-1.20) |
| **Restricted to offspring without own psychiatric disorders** | | | | |
| Unexposed | 235,429 | 27.6 | 1 (Ref) | 1 (Ref) |
| Exposed | 7245 | 28.53 | 1.44 (1.41-1.48) | 1.26 (1.23-1.29) |
| **Restricted to offspring without maternal diagnosis of psychiatric disorders after birth** | | | | |
| Unexposed | 256,112 | 28.45 | 1.00 (ref) | 1.00 (ref) |
| Exposed | 7630 | 28.69 | 1.42 (1.39-1.46) | 1.23 (1.20-1.26) |
| **Additional adjustment for maternal smoking ^c^** | | | | |
| Unexposed | 158,429 | 22.8 | 1 (Ref) | 1 (Ref) |
| Exposed | 8069 | 24.17 | 1.31 (1.28-1.34) | 1.18 (1.15-1.21) |
| **Additional adjustment for maternal pre-pregnancy body-mass index ^d^** | | | | |
| Unexposed | 121,160 | 22.29 | 1 (Ref) | 1 (Ref) |
| Exposed | 6567 | 23.25 | 1.31 (1.27-1.34) | 1.18 (1.15-1.21) |
| **Stratified by maternal family history of CVD before birth** | | | | |
| **Without a record of maternal family history of CVD** | | | | |
| Unexposed | 292,707 | 26.69 | 1.00 (ref) | 1.00 (ref) |
| Exposed | 10,045 | 26.03 | 1.37 (1.34-1.39) | 1.18 (1.16-1.21) |
| **With a maternal family history of CVD** | | | | |
| Unexposed | 4288 | 30.19 | 1.00 (ref) | 1.00 (ref) |
| Exposed | 596 | 31.54 | 1.29 (1.18-1.40) | 1.20 (1.10-1.31) |
| **Stratified by attained age at follow-up** | | | | |
| **Attained age <10 years** | | | | |
| Unexposed | 35,753 | 9.24 | 1.00 (ref) | 1.00 (ref) |
| Exposed | 3229 | 15.86 | 1.72 (1.66–1.78) | 1.23 (1.19-1.28) |
| **10 ≤ Attained age <20 years** | | | | |
| Unexposed | 55,042 | 16.99 | 1.00 (ref) | 1.00 (ref) |
| Exposed | 2671 | 22.53 | 1.42 (1.37-1.48) | 1.18 (1.14-1.23) |
| **20 ≤ Attained age <30 years** | | | | |
| Unexposed | 85,423 | 37.44 | 1.00 (ref) | 1.00 (ref) |
| Exposed | 2531 | 46.99 | 1.27 (1.22-1.32) | 1.20 (1.15-1.25) |
| **30 ≤ Attained age <40 years** | | | | |
| Unexposed | 79,918 | 62.88 | 1.00 (ref) | 1.00 (ref) |
| Exposed | 1688 | 70.54 | 1.13 (1.08-1.19) | 1.15 (1.10-1.21) |
| **Attained age ≥ 40 years** | | | | |
| Unexposed | 40,381 | 90.67 | 1.00 (ref) | 1.00 (ref) |
| Exposed | 522 | 105.03 | 1.21 (1.11-1.32) | 1.19 (1.09-1.29) |
| **Stratified by sex** | | | | |
| Male | | | | |
| Unexposed | 150,903 | 26.39 | 1 (Ref) | 1 (Ref) |
| Exposed | 5559 | 26.79 | 1.41 (1.37-1.44) | 1.20 (1.16-1.23) |
| **Female** | | | | |
| Unexposed | 146,092 | 27.1 | 1 (Ref) | 1 (Ref) |
| Exposed | 5082 | 25.76 | 1.36 (1.32-1.40) | 1.18 (1.15-1.22) |
| **Stratified by birth year** | | | | |
| **Birth year before 2001** | | | | |
| Unexposed | 261,977 | 28.61 | 1 (Ref) | 1 (Ref) |
| Exposed | 6679 | 32.11 | 1.27 (1.24-1.30) | 1.20 (1.17-1.23) |
| **Birth year in 2001 or later** | | | | |
| Unexposed | 35,018 | 17.94 | 1 (Ref) | 1 (Ref) |
| Exposed | 3962 | 20.13 | 1.18 (1.14-1.22) | 1.18 (1.14-1.22) |

Abbreviations: No, number; CVD, cardiovascular disease; HR, hazard ratio; CI, confidence interval.

^a^ Adjusted for offspring’s sex and birth year, maternal country of origin, parity, age, education level, and marital status, family history of cardiovascular diseases prior to the index birth, and hypertensive disorders and diabetes before or during the index pregnancy.

^b^ This analysis was restricted to offspring with register data on father.

^c^ Analysis was restricted to offspring with complete data on maternal smoking.

^d^ Analysis was restricted to offspring with complete data on maternal pre-pregnancy body-mass index

### eTable 5. Hazard ratios and 95% confidence intervals for overall and specific cardiovascular diseases in offspring according to maternal psychiatric disorders in the full cohort (N=4,171,005)

| **Outcome** | **No. of cases** | **Incident rate (10000 person-years)** | **HR (95% CI),**  **Model 1** | **HR (95% CI),**  **Model 2** | **HR (95% CI),**  **Model 3** |
| --- | --- | --- | --- | --- | --- |
| **Overall CVD** |  |  |  |  |  |
| Unexposed | 296995 | 26.73 | 1 (Ref) | 1 (Ref) | 1 (Ref) |
| Exposed | 10641 | 26.28 | 1.38 (1.36-1.41) | 1.19 (1.17-1.22) | 1.19 (1.17-1.21) |
| **IHD** |  |  |  |  |  |
| Unexposed | 5278 | 0.46 | 1 (Ref) | 1 (Ref) | 1 (Ref) |
| Exposed | 117 | 0.28 | 1.32 (1.10-1.59) | 1.26 (1.05-1.52) | 1.26 (1.05-1.52) |
| **AMI** |  |  |  |  |  |
| Unexposed | 3077 | 0.27 | 1 (Ref) | 1 (Ref) | 1 (Ref) |
| Exposed | 61 | 0.15 | 1.28 (0.99-1.65) | 1.22 (0.95-1.58) | 1.22 (0.95-1.58) |
| **Stroke** |  |  |  |  |  |
| Unexposed | 9421 | 0.83 | 1 (Ref) | 1 (Ref) | 1 (Ref) |
| Exposed | 282 | 0.68 | 1.26 (1.12-1.41) | 1.12 (0.99-1.26) | 1.12 (0.99-1.26) |
| **Hemorrhagic stroke** |  |  |  |  |  |
| Unexposed | 2464 | 0.22 | 1 (Ref) | 1 (Ref) | 1 (Ref) |
| Exposed | 92 | 0.22 | 1.44 (1.17-1.78) | 1.27 (1.03-1.58) | 1.27 (1.03-1.58) |
| **Ischemic stroke** |  |  |  |  |  |
| Unexposed | 5399 | 0.47 | 1 (Ref) | 1 (Ref) | 1 (Ref) |
| Exposed | 147 | 0.35 | 1.22 (1.03-1.44) | 1.03 (0.87-1.22) | 1.03 (0.87-1.22) |
| **Heart failure** |  |  |  |  |  |
| Unexposed | 4648 | 0.41 | 1 (Ref) | 1 (Ref) | 1 (Ref) |
| Exposed | 190 | 0.46 | 1.83 (1.58-2.11) | 1.59 (1.37-1.85) | 1.59 (1.37-1.85) |
| **Atrial fibrillation** |  |  |  |  |  |
| Unexposed | 9907 | 0.87 | 1 (Ref) | 1 (Ref) | 1 (Ref) |
| Exposed | 197 | 0.48 | 1.04 (0.90-1.20) | 1.04 (0.90-1.20) | 1.04 (0.90-1.20) |
| **Hypertensive disease** |  |  |  |  |  |
| Unexposed | 19231 | 1.69 | 1 (Ref) | 1 (Ref) | 1 (Ref) |
| Exposed | 554 | 1.34 | 1.38 (1.27-1.50) | 1.22 (1.12-1.33) | 1.22 (1.12-1.32) |
| **Peripheral arterial disease** |  |  |  |  |  |
| Unexposed | 1365 | 0.12 | 1 (Ref) | 1 (Ref) | 1 (Ref) |
| Exposed | 38 | 0.09 | 1.16 (0.84-1.60) | 1.12 (0.81-1.56) | 1.12 (0.81-1.56) |

Model 1 is unadjusted, with the offspring’s attained age as underlying timescale. Model 2 is adjusted for offspring’s sex and birth year, maternal country of origin, parity, age, education, marital status, and family history of cardiovascular diseases prior to the index birth. Model 3 is further adjusted for maternal hypertensive disorders and diabetes before or during the index pregnancy.

Abbreviations: No, number; HR, hazard ratio; CI, confidence interval; CVD, cardiovascular disease; IHD, ischemic heart disease; AMI, acute myocardial infarction.

### eTable 6. Hazard ratios and 95% confidence intervals for overall and specific cardiovascular diseases in offspring according to maternal psychiatric disorders in the cousin comparison cohort (N=1,577,113)

|  | **No. of cases** | **Incident rate (10,000 person-years)** | **HR (95% CI)** | | |
| --- | --- | --- | --- | --- | --- |
|  |  |  | **Model 1** | | **Model 2** |
| **Overall CVD** | | | | | |
| Unexposed | 117,506 | 26.60 | 1 (Ref) | 1 (Ref) | |
| Exposed | 3866 | 26.49 | 1.14 (1.09-1.19) | 1.08 (1.03-1.13) | |
| **IHD** | | | | | |
| Unexposed | 2020 | 0.45 | 1 (Ref) | 1 (Ref) | |
| Exposed | 50 | 0.33 | 1.11 (0.73–1.70) | 1.20 (0.78-1.84) | |
| **AMI** | | | | | |
| Unexposed | 1165 | 0.26 | 1 (Ref) | 1 (Ref) | |
| Exposed | 24 | 0.16 | 0.81 (0.44-1.48) | 0.87 (0.47-1.63) | |
| **Stroke** | | | | | |
| Unexposed | 3676 | 0.81 | 1 (Ref) | 1 (Ref) | |
| Exposed | 96 | 0.64 | 1.15 (0.85-1.56) | 1.14 (0.84-1.55) | |
| **Hemorrhagic stroke** | | | | | |
| Unexposed | 965 | 0.21 | 1 (Ref) | 1 (Ref) | |
| Exposed | 30 | 0.20 | 1.26 (0.72-2.19) | 1.23 (0.70-2.16) | |
| **Ischemic stroke** | | | | | |
| Unexposed | 2095 | 0.46 | 1 (Ref) | 1 (Ref) | |
| Exposed | 52 | 0.35 | 1.17 (0.77–1.77) | 1.13 (0.75-1.72) | |
| **Heart failure** | | | | | |
| Unexposed | 1840 | 0.41 | 1 (Ref) | 1 (Ref) | |
| Exposed | 81 | 0.54 | 1.69 (1.19-2.42) | 1.51 (1.06-2.17) | |
| **Atrial fibrillation** | | | | | |
| Unexposed | 3878 | 0.86 | 1 (Ref) | 1 (Ref) | |
| Exposed | 89 | 0.60 | 1.36 (0.97-1.91) | 1.35 (0.97-1.90) | |
| **Hypertensive disease** | | | | | |
| Unexposed | 7545 | 1.67 | 1 (Ref) | 1 (Ref) | |
| Exposed | 205 | 1.37 | 1.12 (0.91-1.39) | 1.07 (0.86-1.32) | |
| **Peripheral arterial disease** | | | | | |
| Unexposed | 504 | 0.11 | 1 (Ref) | 1 (Ref) | |
| Exposed | 10 | 0.07 | 0.60 (0.27-1.38) | 0.68 (0.29-1.56) | |

Model 1 is unadjusted, with the offspring’s attained age as underlying timescale. Model 2 is adjusted for offspring’s sex and birth year, maternal country of origin, parity, age, education level, and marital status, and hypertensive disorders and diabetes before or during the index pregnancy.

Abbreviations: No, number; HR, hazard ratio; CI, confidence interval; CVD, cardiovascular disease; IHD, ischemic heart disease; AMI, acute myocardial infarction.

### eTable 7. Hazard ratios and 95% confidence intervals for overall cardiovascular diseases in offspring according to specific maternal psychiatric disorders in the full cohort (N=4,171,005)

| **Exposure** | **No. of cases** | **Incident rate (10000 person-years)** | **HR (95% CI),**  **Model 1** | **HR (95% CI),**  **Model 2** | **HR (95% CI),**  **Model 3** |
| --- | --- | --- | --- | --- | --- |
| Organic psychoses | 277 | 29.6 | 1.21 (1.08-1.37) | 1.17 (1.04-1.32) | 1.17 (1.04-1.31) |
| Schizophrenia and other non-affective psychoses | 633 | 27.12 | 1.15 (1.06-1.24) | 1.10 (1.01-1.18) | 1.09 (1.01-1.18) |
| Affective disorders | 1660 | 21.13 | 1.65 (1.57-1.73) | 1.17 (1.11-1.23) | 1.17 (1.11-1.23) |
| Neurotic disorders | 4417 | 29.13 | 1.37 (1.33-1.41) | 1.25 (1.21-1.28) | 1.25 (1.21-1.28) |
| Stress-related disorders | 1960 | 24.36 | 1.53 (1.46-1.59) | 1.20 (1.14-1.25) | 1.19 (1.14-1.25) |
| Personality disorder | 1104 | 28.82 | 1.38 (1.30-1.47) | 1.23 (1.16-1.31) | 1.23 (1.16-1.31) |
| Attention deficit hyperactivity disorders | 116 | 17.89 | 1.83 (1.52-2.19) | 1.09 (0.90-1.30) | 1.08 (0.90-1.30) |
| Autism spectrum disorders | 24 | 14.87 | 1.41 (0.95-2.11) | 0.87 (0.58-1.30) | 0.87 (0.58-1.30) |
| Disorders related to psychoactive substance use | 2150 | 26.36 | 1.46 (1.40-1.52) | 1.21 (1.16-1.26) | 1.21 (1.16-1.26) |
| Intellectual disability | 177 | 29.62 | 1.38 (1.19-1.60) | 1.22 (1.05-1.41) | 1.21 (1.05-1.40) |
| Somatoform disorders | 457 | 28.8 | 1.64 (1.49-1.79) | 1.35 (1.23-1.48) | 1.35 (1.23-1.48) |

Model 1 is unadjusted, with the offspring’s attained age as underlying timescale. Model 2 is adjusted for offspring’s sex and birth year, maternal country of origin, parity, age, education, marital status, and family history of cardiovascular diseases prior to the index birth. Model 3 is further adjusted for maternal hypertensive disorders and diabetes before or during the index pregnancy.

Abbreviations: No., number, HR, hazard ratio; CI, confidence interval; CVD, cardiovascular disease; IHD, ischemic heart disease; AMI, acute myocardial infarction.

### eTable 8. Hazard ratios and 95% confidence intervals for overall cardiovascular diseases in offspring according to specific maternal psychiatric disorders in the cousin comparison cohort (N=1,577,113)

| **Exposure** | **No. of cases** | **Incident rate (10,000 person-years)** | **HR (95% CI)** | |
| --- | --- | --- | --- | --- |
|  |  |  | **Model 1** | **Model 2** |
| Organic psychoses | 100 | 28.80 | 0.92 (0.70-1.23) | 0.87 (0.66-1.16) |
| Schizophrenia and other non-affective psychoses | 223 | 26.29 | 0.97 (0.80-1.16) | 0.91 (0.75-1.09) |
| Affective disorders | 532 | 20.53 | 1.12 (0.99-1.26) | 1.02 (0.91-1.16) |
| Neurotic disorders | 1588 | 28.76 | 1.17 (1.08-1.26) | 1.11 (1.03-1.19) |
| Stress-related disorders | 714 | 25.98 | 1.17 (1.05-1.31) | 1.09 (0.98-1.21) |
| Personality disorder | 401 | 29.39 | 1.16 (1.01-1.34) | 1.09 (0.94-1.26) |
| Attention deficit hyperactivity disorders | 26 | 16.32 | 0.87 (0.47-1.58) | 0.77 (0.42-1.41) |
| Autism spectrum disorders | <10 | 7.88 | 0.73 (0.15-3.60) | 0.66 (0.14-3.24) |
| Disorders related to psychoactive substance use | 758 | 26.89 | 1.14 (1.03-1.28) | 1.06 (0.95-1.19) |
| Intellectual disability | 65 | 28.30 | 0.90 (0.64-1.27) | 0.86 (0.61-1.21) |
| Somatoform disorders | 167 | 29.56 | 1.63 (1.28-2.07) | 1.55 (1.23-1.97) |

Model 1 is unadjusted, with the offspring’s attained age as underlying timescale. Model 2 is adjusted for offspring’s sex and birth year, maternal country of origin, parity, age, education level, and marital status, and hypertensive disorders and diabetes before or during the index pregnancy.

Abbreviations: No, number; HR, hazard ratio; CI, confidence interval; CVD, cardiovascular disease; IHD, ischemic heart disease; AMI, acute myocardial infarction.

### eTable 9. Hazard ratios and 95% confidence intervals for overall and specific cardiovascular diseases in offspring according to maternal psychiatric disorders in the propensity-score-matched sub-cohort

|  | **No. of cases** | **Incident rate (10,000 person-years)** | **HR (95% CI)** | | |
| --- | --- | --- | --- | --- | --- |
|  |  |  | **Model 1 ^a^** | | **Model 2 ^a^** |
| **Overall CVD** | | | | | |
| Unexposed | 9102 | 22.39 | 1 (Ref) | 1 (Ref) | |
| Exposed | 10,641 | 26.28 | 1.18 (1.15-1.21) | 1.18 (1.15-1.21) | |
| **IHD** | | | | | |
| Unexposed | 93 | 0.22 | 1 (Ref) | 1 (Ref) | |
| Exposed | 117 | 0.28 | 1.26 (0.96-1.66) | 1.25 (0.95-1.64) | |
| **AMI** | | | | | |
| Unexposed | 49 | 0.12 | 1 (Ref) | 1 (Ref) | |
| Exposed | 61 | 0.15 | 1.25 (0.86-1.82) | 1.24 (0.85-1.80) | |
| **Stroke** | | | | | |
| Unexposed | 280 | 0.67 | 1 (Ref) | 1 (Ref) | |
| Exposed | 282 | 0.68 | 1.01 (0.86-1.19) | 1.01 (0.86-1.19) | |
| **Hemorrhagic stroke** | | | | | |
| Unexposed | 77 | 0.19 | 1 (Ref) | 1 (Ref) | |
| Exposed | 92 | 0.22 | 1.20 (0.88-1.62) | 1.19 (0.88-1.62) | |
| **Ischemic stroke** | | | | | |
| Unexposed | 157 | 0.38 | 1 (Ref) | 1 (Ref) | |
| Exposed | 147 | 0.35 | 0.94 (0.75-1.17) | 0.94 (0.75-1.18) | |
| **Heart failure** | | | | | |
| Unexposed | 135 | 0.41 | 1 (Ref) | 1 (Ref) | |
| Exposed | 190 | 0.46 | 1.42 (1.13-1.76) | 1.41 (1.13-1.76) | |
| **Atrial fibrillation** | | | | | |
| Unexposed | 218 | 0.53 | 1 (Ref) | 1 (Ref) | |
| Exposed | 197 | 0.48 | 0.91 (0.75-1.10) | 0.90 (0.74-1.09) | |
| **Hypertensive disease** | | | | | |
| Unexposed | 431 | 1.04 | 1 (Ref) | 1 (Ref) | |
| Exposed | 554 | 1.34 | 1.29 (1.14-1.46) | 1.28 (1.13-1.46) | |
| **Peripheral arterial disease** | | | | | |
| Unexposed | 22 | 0.05 | 1 (Ref) | 1 (Ref) | |
| Exposed | 38 | 0.09 | 1.73 (1.02-2.93) | 1.72 (1.02-2.91) | |

Model 1 is unadjusted, with the offspring’s attained age as underlying timescale. Model 2 is adjusted for offspring’s sex and birth year, maternal country of origin, parity, age, education level, and marital status, family history of cardiovascular diseases prior to the index birth, and hypertensive disorders and diabetes before or during the index pregnancy.

Abbreviations: No, number; HR, hazard ratio; CI, confidence interval; CVD, cardiovascular disease; IHD, ischemic heart disease; AMI, acute myocardial infarction.

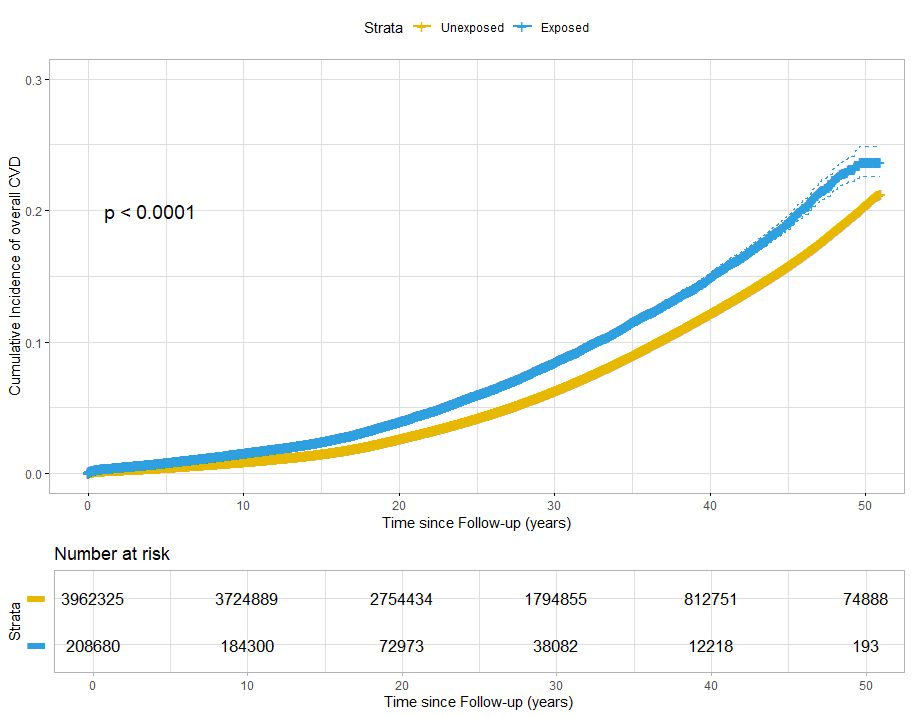

### eFigure 1. Cumulative incidence of overall cardiovascular disease in offspring according to maternal psychiatric disorders

CVD, cardiovascular disease.

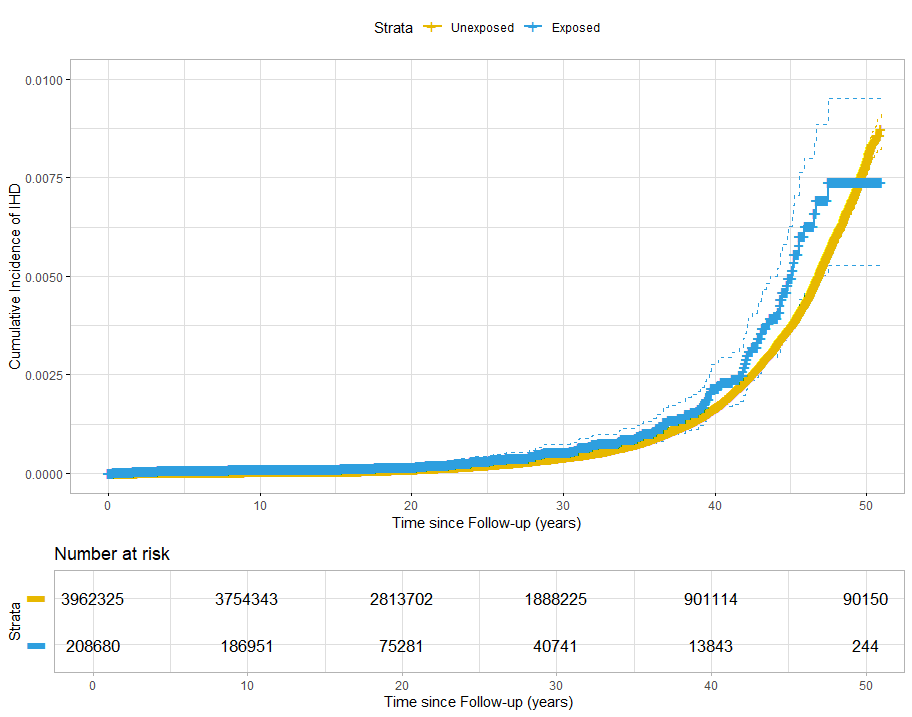

### eFigure 2. Cumulative incidence of ischemic heart disease in offspring according to maternal psychiatric disorders

IHD, ischemic heart disease.

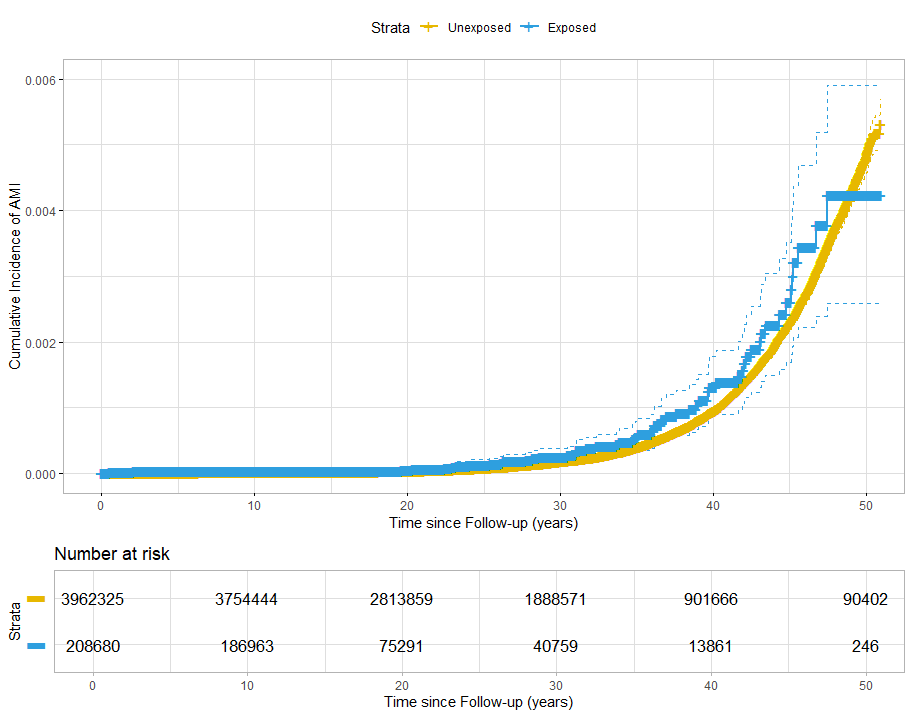

### eFigure 3. Cumulative incidence of acute myocardial infarction in offspring according to maternal psychiatric disorders

AMI, acute myocardial infarction.

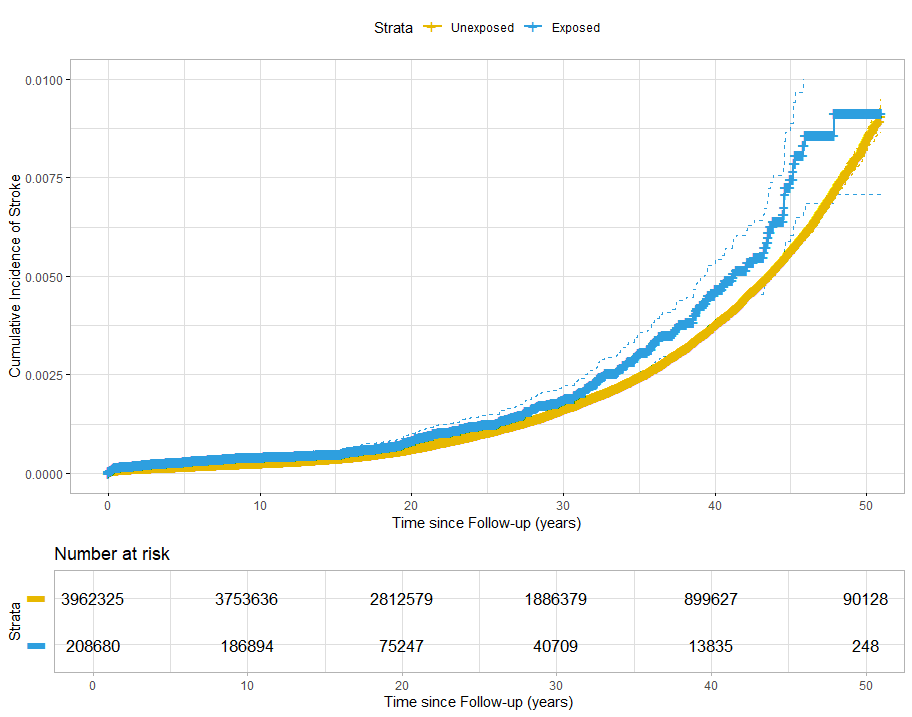

### eFigure 4. Cumulative incidence of stroke in offspring according to maternal psychiatric disorders

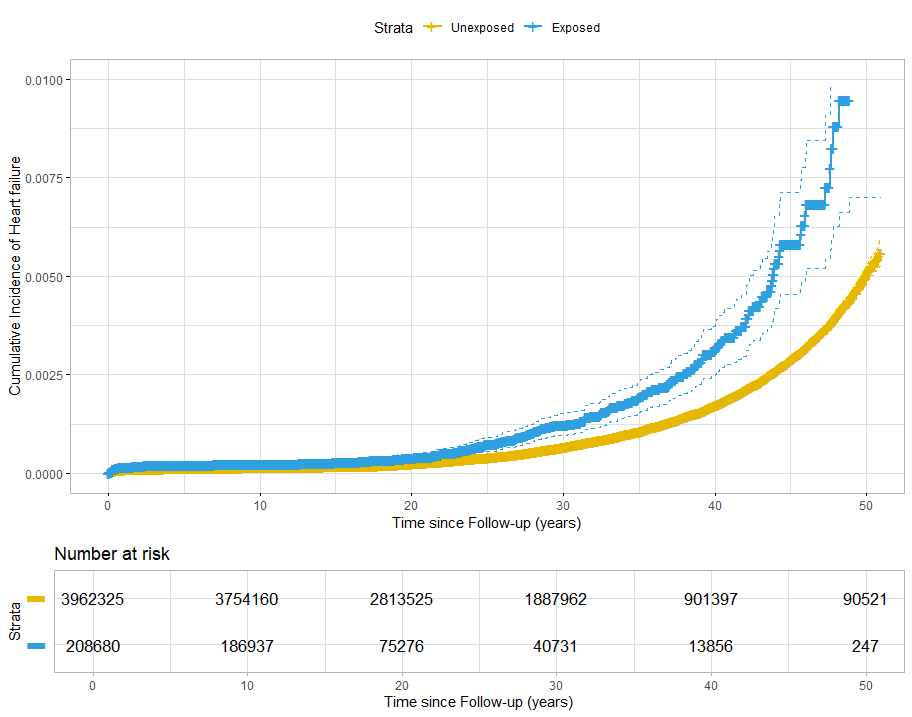

### eFigure 5. Cumulative incidence of heart failure in offspring according to maternal psychiatric disorders

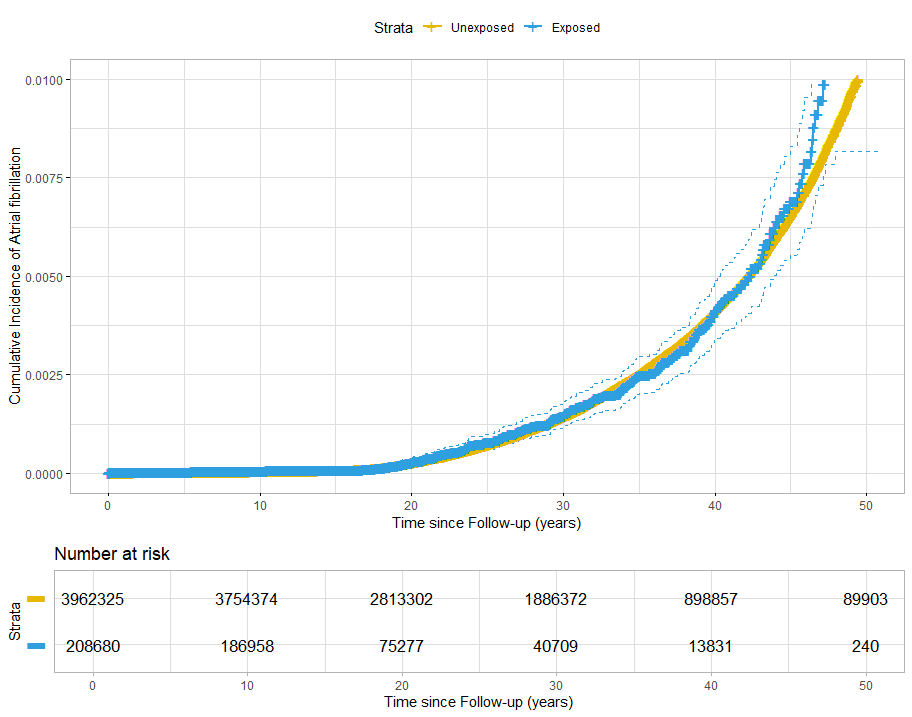

### eFigure 6. Cumulative incidence of atrial fibrillation in offspring according to maternal psychiatric disorders

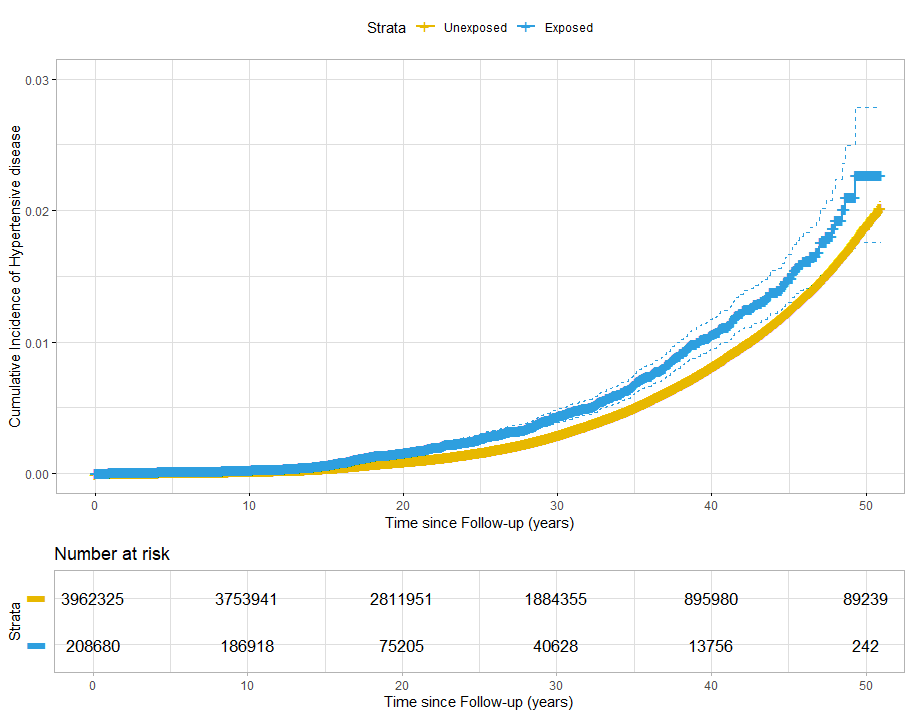

### eFigure 7. Cumulative incidence of hypertensive disease in offspring according to maternal psychiatric disorders

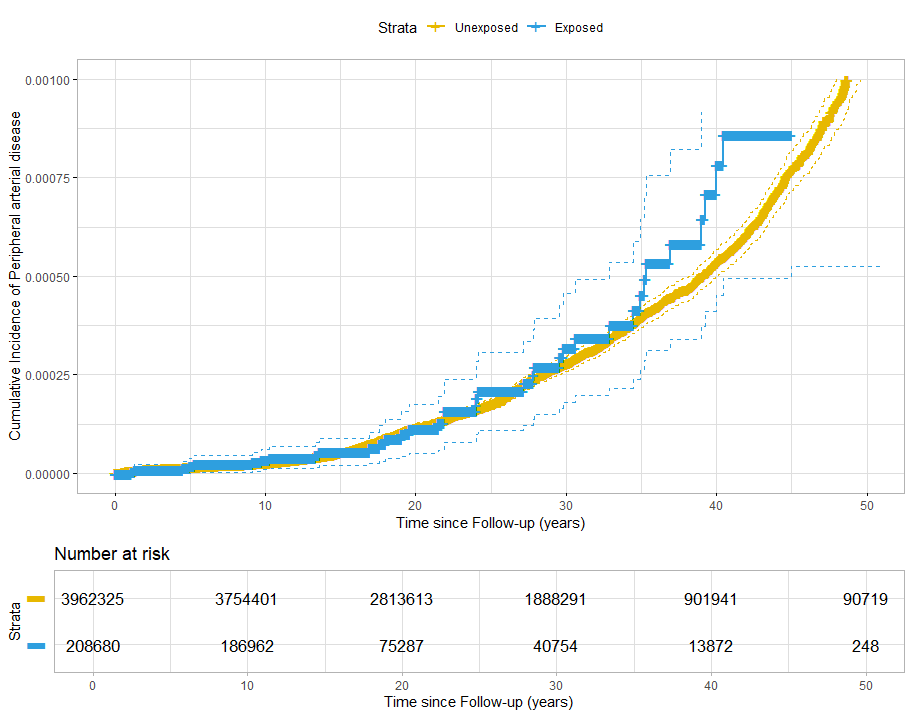

### eFigure 8. Cumulative incidence of peripheral arterial disease in offspring according to maternal psychiatric disorders

### eAppendix 1. Covariates and their source

We selected the following covariates that are associated with maternal psychiatric disorders or offspring cardiovascular disease or both, based on prior knowledge.

Maternal age at delivery (≤19, 20-24, 25-29, 30-34, or ≥35 years), parity (1, 2, ≤3), body-mass index (BMI) (<18.5, 18.5-24.9, 25.0-29.9, or ≥30 kg/m^2^) and smoking in early pregnancy (the latter two available since 1982 in Sweden) were extracted from the Medical Birth Register. Maternal country of origin (Sweden or other countries) and marital status (married/registered partnership or not) at the time of index birth was extracted from the Swedish Total Population Register. Maternal highest level of education (primary and lower secondary, upper secondary, bachelor, or higher) at index birth was obtained from the Register of Education. Information on maternal hypertensive disease and diabetes (International Classification of Disease codes in Supplementary Table S2) before or during the index pregnancy were extracted from the Medica Birth Register and the Patient Register. Maternal family history of cardiovascular disease before the index birth was retrieved from the Patient Register.

Paternal psychiatric disorders before the index birth was retrieved from the Patient Register using the International Classification of Disease codes in Supplementary Table S2.

Offspring characteristics, i.e., birth year (categorised as 1973-1978, 1979-1984, 1985-1990, 1991-1996, 1997-2002, 2003-2008, or 2009-2014), sex (male or female), gestational age and birth weight were retrieved from the Medical Birth Register. The diagnosis of congenital heart defects during the follow-up period was extracted from the Patient Register using the International Classification of Disease codes presented in Supplementary Table 2.
